# Mobility-Guided Modeling of the COVID-19 Pandemic in Metro Manila

**DOI:** 10.1101/2020.05.26.20111617

**Authors:** Bernhard Egwolf, O.P. Nicanor Austriaco

## Abstract

COVID-19 is a novel respiratory disease first identified in Wuhan, China, that is caused by the novel coronavirus, SARS-CoV-2. To better understand the dynamics of the COVID-19 pandemic in the Philippines, we have used real-time mobility data to modify the DELPHI Epidemiological Model recently developed at M.I.T., and to simulate the pandemic in Metro Manila. We have chosen to focus on the National Capital Region, not only because it is the nation’s demographic heart where over a tenth of the country’s population live, but also because it has been the epidemiological epicenter of the Philippine pandemic. Our UST CoV-2 model suggests that the government-imposed enhanced community quarantine (ECQ) has successfully limited the spread of the pandemic. It is clear that the initial wave of the pandemic is flattening, though suppression of viral spread has been delayed by the local pandemics in the City of Manila and Quezon City. Our data also reveals that replacing the ECQ with a General Community Quarantine (GCQ) will increase the forecasted number of deaths in the nation’s capital unless rigorous tracing and testing can be implemented to prevent a second wave of the pandemic.

## INTRODUCTION

COVID-19 is a novel respiratory disease first identified in Wuhan, China, that is caused by the coronavirus, SARS-CoV-2 (Guan et al., 2020; Xie et al., 2020; Zhu et al., 2020). With widespread human-to-human transmission, the virus is highly contagious, and the COVID-19 pandemic is now of global concern (Burki, 2020; Paules et al., 2020).

On January 30, 2020, the Department of Health (DOH) of the Philippines reported its first case of COVID-19 in the country. She was a 38-year-old female Chinese national who had travelled to the Philippines from Wuhan via Hong Kong. The first case of local transmission was confirmed on March 7, 2020, with the first death due to local transmission reported on March 11, 2020. As of May 21, 2020, there have been 13,434 confirmed cases and 846 deaths from COVID-19 reported by the DOH throughout the archipelago.

To contain the COVID-19 pandemic, the national government of the Philippines imposed an Enhanced Community Quarantine (ECQ) on the National Capital Region (NCR) of the country, also known as Metro Manila, on March 15, 2020, which was extended to the entire island of Luzon and its 55 million inhabitants the following day. The ECQ imposed a strict stay-at-home order, banned all public gatherings, suspended all mass public transportation, and closed all non-essential business establishments. On April 24, 2020, the ECQ was extended in the NCR and several other provinces and municipalities that were considered high-risk for COVID-19. On May 15, 2020, Metro Manila and several surrounding regions were placed on a Modified Enhanced Community Quarantine (MECQ) that is to stay in place until May 31, 2020. At that time, if conditions permit, the National Capital Region is expected to move to a more relaxed General Community Quarantine (GCQ).

Numerous mathematical models have been developed to understand the global COVID-19 pandemic (Adam, 2020; Currie et al., 2020; Holmdahl & Buckee, 2020). To better understand the dynamics of the local pandemic in the Philippines, we have used real-time mobility data to adapt the DELPHI Epidemiological Model recently developed at M.I.T. (Bertsimas et al., 2020), to the pandemic in Metro Manila. The DELPHI model is a standard SEIR model with additional features, including under-detection and differentiated government intervention, that are particular helpful for modeling this current pandemic. Finally, it uses a machine learning algorithm to determine the best-fit epidemiological parameters from the historical data of death counts and detected cases. The current SEIR models for the COVID-19 pandemic in the Philippines do not have these enhancements (David et al., 2020).

We have chosen to focus on the National Capital Region of the Philippines, not only because it is the nation’s demographic heart where over a tenth of the country’s population live, but also because it has been, by far, the epidemiological epicenter of the Philippine pandemic: As of May 21, 2020, 8,659 cases (64% of the country’s total) and 621 deaths (73%) have been reported in Metro Manila.

Our UST CoV-2 mobility-guided enhanced SEIR model suggests that the lockdown measures of the ECQ have successfully limited the spread of the pandemic in the NCR. It is clear that the initial wave of the pandemic is flattening, though suppression of viral spread has been delayed by the local pandemics in Quezon City and the City of Manila. Our model also reveals that releasing these measures will increase the forecasted number of deaths in the nation’s capital unless rigorous tracing and testing can be implemented to prevent a second wave of the pandemic.

## MODELING METHODS

The DELPHI (Differential Equations Leads to Predictions of Hospitalizations and Infections) Epidemiological Model recently developed at M.I.T. is a compartmental model that is based on the successful SEIR models that have been used to simulate numerous past epidemics (Bertsimas et al., 2020). However, it also accounts for the underdetection of cases, which is particularly important when a community is unable to adequately test for the disease during the pandemic, and for possible societal-governmental responses to contain spread of the contagion. Both enhancements are essential for robust modeling the current COVID-19 pandemic.

To model the underdetection of cases, the DELPHI model separates the target population into eleven possible categories at the onset of the pandemic, including undetected recovered (AR) and undetected dead (AD) states, which are novel buckets not included in classic SEIR models. It also includes a parameter, p_d_, for the percentage of infection cases detected that allows the model to incorporate varying testing capacities for the target population.

To model the societal-government response, the DELPHI model includes a multiplier for the rate of infection in the form of a smooth parametric nonlinear arctan function:

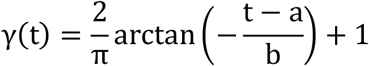

Parameters *a* and *b* can be varied to model a wide range of policies including social distancing, stay-at-home policies, and lockdowns.

Finally, in the original DELPHI model, key parameters for COVID-19 were fixed using a metanalysis conducted by the M.I.T. COVIDAnalytics team, and epidemiological parameters were fitted to historical death counts and detected cases using a machine learning algorithm that best fits these parameters to the observed data.

To simulate the COVID-19 pandemic in Metro Manila, we modified the DELPHI model in four ways:

First, real-time mobility data obtained from Apple, Inc. for driving, walking, and transit on May 21, 2020 (https://www.apple.com/covid19/mobility), revealed that the lockdown in Metro Manila that began on March 15, 2020, resulted in a dramatic and near-immediate drop in mobility over the course of several days. A graph of the changes in mobility in the National Capital Region obtained by first averaging the three mobility data sets provided by Apple, Inc. – driving, walking, and transit – for each date, and then calculating a seven-day running average of these means is shown in Supplemental Figure 1. Therefore, to better simulate the initial period of the local pandemic in Metro Manila, we replaced the original smooth parametric nonlinear arctan function adopted by the M.I.T. team, with an almost step-like function that would be fitted to the historical COVID-19 pandemic data from the Philippine Department of Health for the National Capital Region using a modified machine learning algorithm.

Our replacement is a function γ(t) which can change fast between constant values. The value of γ(t) is set to 1 at the beginning of the COVID-19 outbreak. Two days before the optimized median day of action *t_a_* the function γ(t) starts dropping rapidly to the optimized level of action *L_a_*. This reduces the effective reproduction number R(t) = α γ(t)/r_d_, where a is the infection rate, and r_d_ is the detection rate, as expected during community quarantine. The level of action *L_a_* is reached two days after t_a_. During this four day transition the γ(t) function connects the two constant levels with a cubic spline:

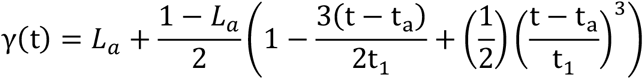

for t_a_-t_1_ < t < t_a_ + t_1_ with the transition time t_1_ = 2 days. The γ(t) function is equal to *L_a_* during the community quarantine. In our view, this novel multiplier developed with mobility data in mind allows us to better simulate the dramatic lowering of the rate of infection that is expected during the initial days of the ECQ because of the dramatic decrease in mobility. Human mobility proxies have been used in the past to understand the spread of infectious diseases including COVID-19 (Bryant & Elofsson, 2020; Muller et al., 2020; Tizzoni et al., 2014)

Next, the Apple Mobility data revealed that the mobility of communities in Europe that were put into relatively lenient lockdowns recovered linearly from the minimum achieved immediately after quarantines were established. Three representative curves from Austria, Germany, and Norway, obtained as above, by first averaging the three mobility data sets provided by Apple, Inc. – driving, walking, and transit – for each date, and then calculating a seven-day running average of these means, are shown in Supplemental Figure 2. It is likely that the shape of this recovery reflects both the gradual easing of the lockdowns and the hesitant movements of a quarantined population that remains wary of the disease. We therefore chose to model the lifting of the community quarantine in Metro Manila with a smooth cubic spline interpolation function that allows us to transition the multiplier of the infection rate from the lockdown level, *L*_a_, to a higher level, between L_a_ and 1:

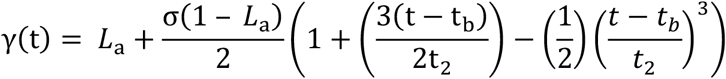

for *t_b_* − *t*_2_ < t < *t*_b_ + *t*_2_. We considered two scenarios: A smooth increase with a transition time *t*_2_ of 7 days, where *t*_b_ corresponds to June 7, 2020, and a very smooth increase with a transition time *t*_2_ of 15 days, where *t*_b_ corresponds to June 15, 2020. The γ(t) function is equal to the constant *L*_a_ + σ (1 − *L*_a_) after this transition. In order to study the effect of different σ values, σ was increased from 0 to 1 in steps of 0.1. A value of 0 for σ means that γ(t) stays at the level *L*_a_ associated with the original lockdown, whereas a value of 1 for σ means that γ(t) returns to the initial value 1. We eventually chose to focus our analysis on a two-week period of transition (*t*_2_ is 7 days) from the ECQ to GCQ because we believe that it would take two weeks for people to adjust to the new more-relaxed quarantine.

Third, we replaced the original machine learning algorithm in the DELPHI model. The variables of the modified model used in this study were optimized by minimizing a function *f*(*a*, *t*_a_, *L*_a_, *r*_dth_, *p*_dth_, *k*_1_, *k*_2_) of the variables, while respecting the upper and lower bounds specified for them. This function is a measure of the distance between the predictions of the DELPHI model and the historical data of death counts and detected cases. The function was minimized with the aid of the dual_annealing algorithm of the Python module SciPy. To reduce the number of variables which need to be optimized, we fixed the infection rate *a* to 2.2*r*_d_. This corresponds to a basic reproduction number *R*_0_ = a/*r*_d_ of 2.2, which is a published estimate for the R_0_ for SARS-CoV-2 (Li et al., 2020). The value of *a* was fixed by specifying upper (2.2*r*_d_ + 0.000000001/days) and lower (2.2*r*_d_ − 0.000000001/days) bounds which are just slightly above and below 2.2*r*_d_.

Finally, we specified several of the epidemiological parameters of the original DELPHI model to better reflect the local pandemic in Metro Manila. The values for the clinical and epidemiological parameters used in our model are as follows:

| MODEL PARAMETER | VALUE | SOURCE |
| --- | --- | --- |
| Median time to leave incubation | 5d | (Bertsimas et al., 2020) |
| Median time to recovery without hospitalization | 10d | (Bertsimas et al., 2020) |
| Median time to detection | 2d | (Bertsimas et al., 2020) |
| Median time to recovery in hospital. | 15d | (Bertsimas et al., 2020) |
| Ratio of recoveries to deaths | 6:1 | worldometers.info |
| $p_d$ for Philippines<br>(% infected cases detected) | 0.02 | (Bommer & Vollmer, 2020) |
| $p_h$ for Metro Manila<br>(% detected cases hospitalized) | 0.10 | doh.gov.ph/covid19tracker |
| Population of Metro Manila | 13,923,452 | worldpopulationreview.com |

Additional parameter values and the initial conditions for our modeling efforts are provided in the Supplementary Information.

All historical data for the pandemic in the National Capital Region was obtained from the COVID-19 Tracker website maintained by the Department of Health of the Philippines (https://ncovtracker.doh.gov.ph/). We have noticed that this historical data set is constantly being revised to reflect the results of ongoing validation tests. Our model used the confirmed values that were available on May 21, 2020, to generate the results described in this paper. Data from New York City was obtained from the website of the New York City Department of Health (https://www1.nyc.gov/site/doh/covid/covid-19-data.page).

## RESULTS & DISCUSSION

### Impact of the Enhanced Community Quarantine (ECQ) in Metro Manila

There is evidence that social distancing practices and community-wide lockdowns have been effective at blunting the COVID-19 pandemic and preventing health systems from being overwhelmed in different countries around the world (Ainslie et al., 2020; Ferguson et al., 2020; Giordano et al., 2020; Prem et al., 2020; Wells et al., 2020). As shown in Figure 1, our UST CoV-2 Model suggests that the government-imposed enhanced community quarantine (ECQ) has also successfully limited the impact of the pandemic in Metro Manila by significantly lowering the total number of COVID-19 cases and the total number of deaths.

**Figure 1:**
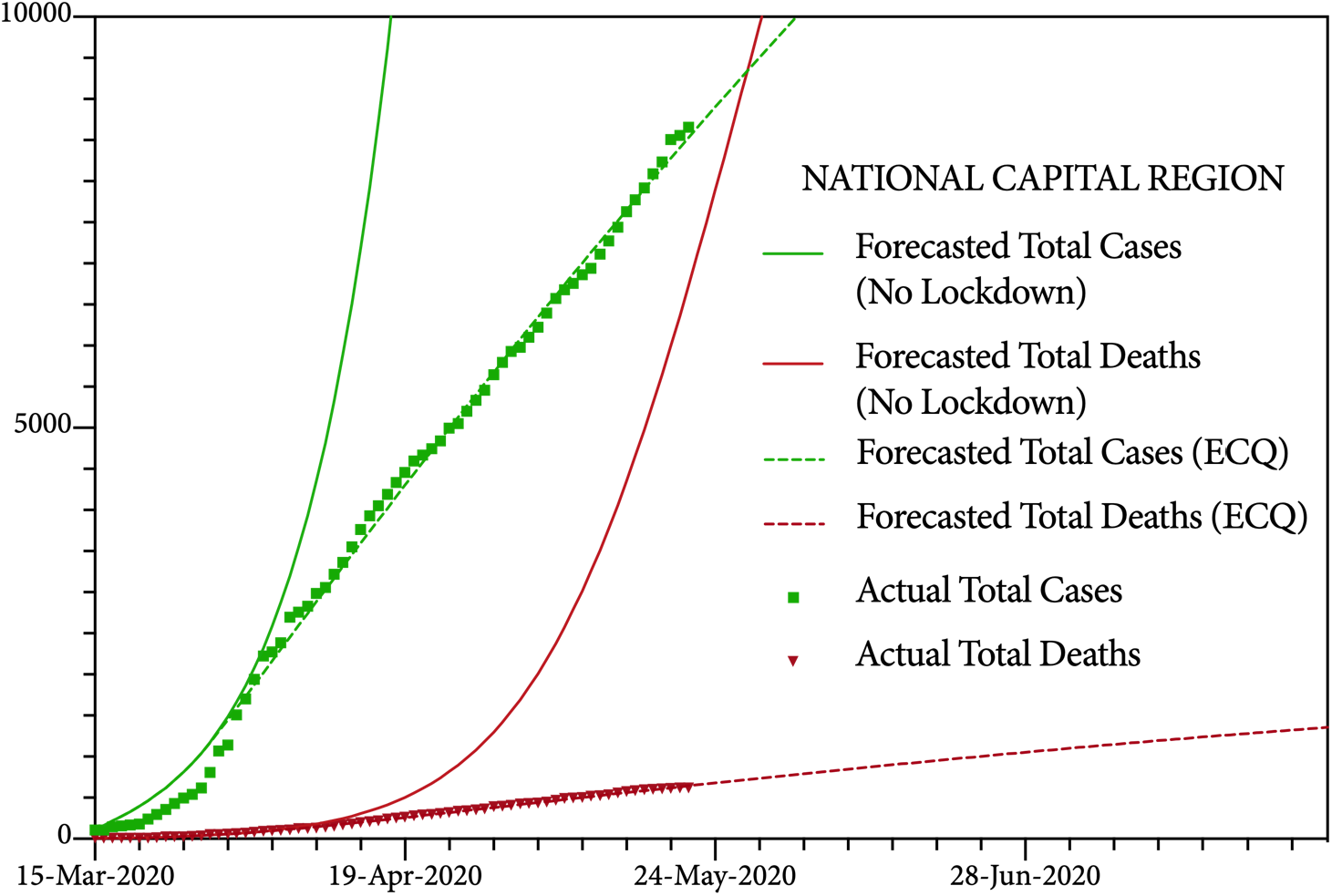
Modeling the Impact of the Enhanced Community Quarantine (ECQ) on the COVID-19 Pandemic in Metro Manila. Forecasted estimates of total COVID-19 cases and total deaths for the National Capital Region without a lockdown and with the Enhanced Community Quarantine compared to the data provided by the Department of Health of the Philippines.

As a point of comparison, Metro Manila and New York City, two metropolitan areas with comparable populations, recorded their first deaths from community spread of COVID-19 on March 11, 2020, and March 13, 2020, respectively. However, Metro Manila entered lockdown on March 15, 2020, while New York City waited one more week to enter its lockdown on March 20, 2020. As of May 21, 2020, Metro Manila reported 621 total deaths from COVID-19 while New York City confirmed 16,232 total deaths. It appears that the early implementation of the ECQ has been able to spare the lives of thousands of residents of the NCR.

Our results also suggest that the first wave of the pandemic is receding. As shown in Figure 2, the number of forecasted active cases peaked in early May and is gradually declining. Currently, there is no evidence for a second wave of the pandemic. Note that we decided not to compare this modeled curve of forecasted active cases with the data for active cases provided by the DOH of the Philippines because we believe that the number of recovered cases is under-reported in this data set: There are cases of COVID-19 patients who were listed as having mild symptoms whose health status remains unknown two months after their initial diagnosis. This is medically inexplicable, and we believe that it is better explained by gaps in the data collection and reporting process during this time of pandemic.

**Figure 2:**
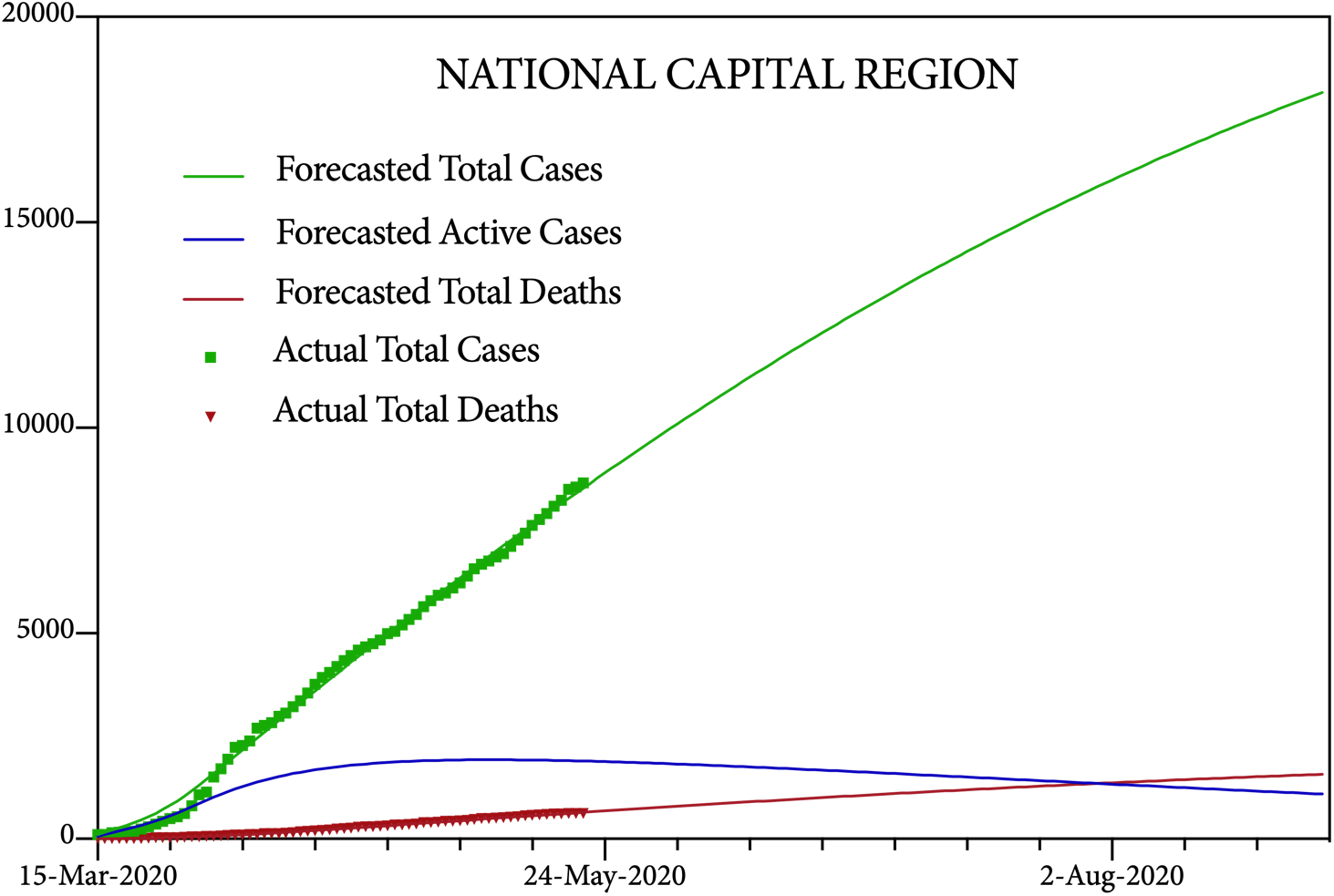
Modeling the COVID-19 Pandemic under Enhanced Community Quarantine (ECQ) in Metro Manila. Forecasted estimates of total COVID-19 cases, total active cases, and total deaths for the National Capital Region with the Enhanced Community Quarantine compared to the data provided by the Department of Health of the Philippines.

Though the curve for the pandemic in the NCR is flattening gradually, the flattening is not dramatic. In fact, it is clear that the ECQ is struggling to suppress the pandemic, i.e., to drive the number of infected cases down to zero. At this rate, the model predicts that the total number of cases in Metro Manila will continue to gradually increase and will not plateau for many months, though the forecasted number of active cases will also be decreasing. If the current quarantine measures are maintained, the forecasted number of active cases of COVID-19 will not fall below 1,000 cases until early September.

To uncover possible reasons for this less-than-dramatic slowing of viral spread, we modeled the local pandemics in the five cities within Metro Manila that have the largest numbers of cases of COVID-19. These are Quezon City and the cities of Makati, Mandaluyong, Manila, and Parañaque. These five cities together represent over half of the total number of COVID-19 cases in the NCR.

Our modeling reveals that the efficacy of efforts to suppress the pandemic have varied widely throughout the NCR. As shown in Figure 3, cities like Makati, Mandaluyong, and Parañaque have been more successful at suppressing the community spread of COVID-19 than Manila or Quezon City. If the current quarantine measures are maintained, the total number of cases is forecasted to plateau by mid-July in Mandaluyong, by early August in Makati, and by September in Parañaque. However, the same cannot be said for the latter two cities where the total number of cases is forecasted to gradually keep increasing for many months, though the forecasted number of active cases will also be decreasing.

**Figure 3:**
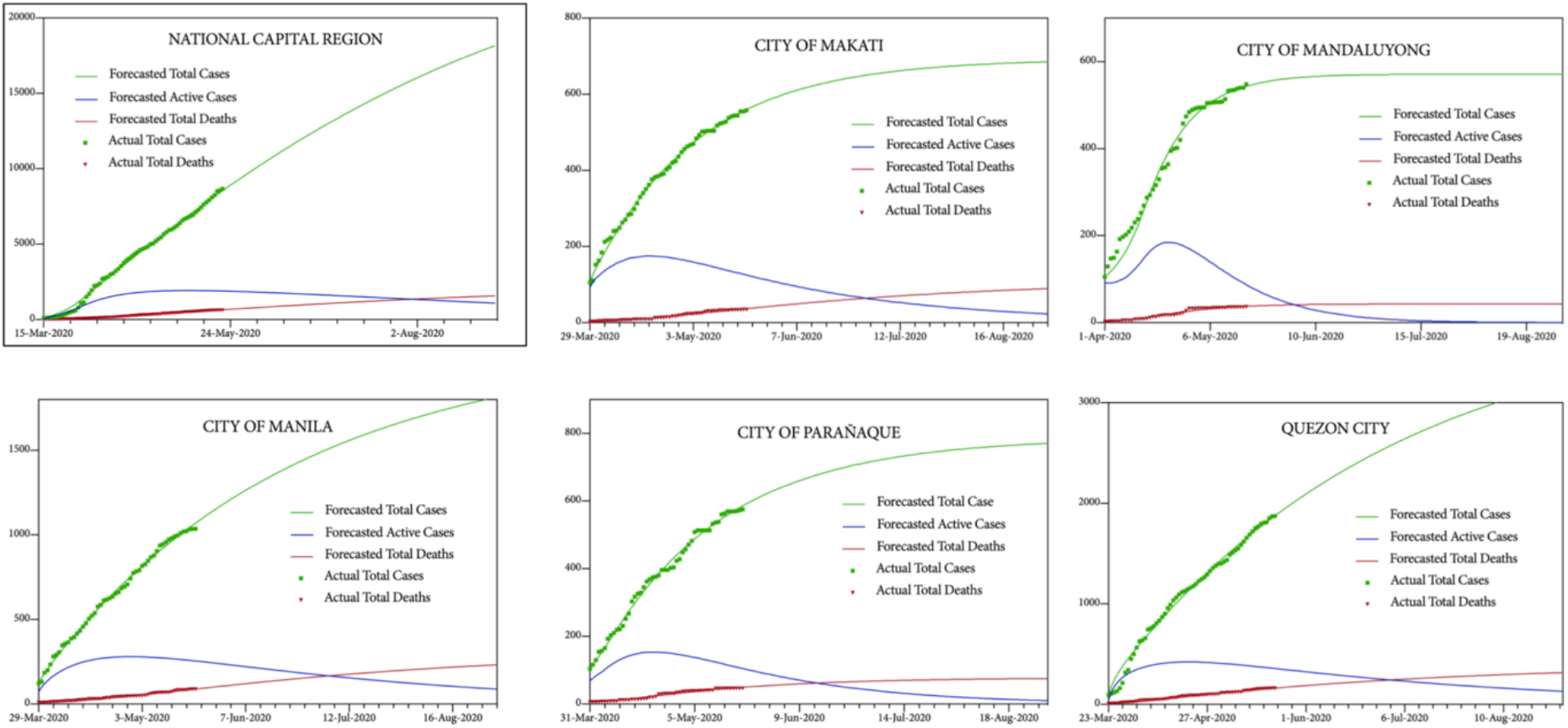
Modeling the COVID-19 Pandemic under Enhanced Community Quarantine (ECQ) in Individual Component Cities of Metro Manila. Forecasted estimates of total cases, total active cases, and total deaths for the National Capital Region and the five component cities with the highest recorded cases of COVID-19 with the Enhanced Community Quarantine compared to the data provided by the Department of Health of the Philippines.

It is not clear why the ECQ has not been as effective in both the City of Manila and Quezon City as it has been in their neighboring municipalities. They are not the top two most dense cities in the NCR so it is unlikely to be explained by appealing to population density alone. Nonetheless, both these urban areas have experienced a several-fold higher number of infections than the other component cities in the capital region. Thus, it is not unreasonable to posit that the dynamics of the COVID-19 pandemic in Metro Manila are being driven by both local pandemics. It suggests that public health authorities should increase their efforts to contain the pandemic in the NCR in these two cities.

### Impact of Easing the Enhanced Community Quarantine (ECQ) in Metro Manila

To model the impact of replacing the Enhanced Community Quarantine (ECQ) with the General Community Quarantine (GCQ), we reran our model assuming that the ECQ would be eased on June 1, 2020, and that it would take two weeks for people to adjust to the new more-relaxed quarantine. As explained above, we had observed that the mobilities of communities released from lockdowns in Europe recovered in a linear fashion. Therefore, we chose to model the easing of the ECQ in Metro Manila with a smooth cubic spline interpolation function that would increase the modeled infection rate in a relatively linear rate once the strict community lockdown is lifted. Since the proposed GCQ (http://www.covid19.gov.ph/ecq-gcq-guidelines/) will seek to limit the mobility of the relatively young (<21 years old) and the relatively old (>60 years old), two populations who together constitute about half of the number of the Filipino people nation-wide (https://www.populationpyramid.net/Philippines/2019/), we interrogated the effect on the pandemic of increasing the infection rate by half of the amount that it decreased during the ECQ, i.e., a post-ECQ recovery rate of 50% in the infection rate that would mirror the expected post-ECQ recovery rate of 50% in the mobility of the residents of the NCR.

As shown in Figure 4, it is clear that replacing the ECQ with a GCQ will increase the forecasted number of infected cases and deaths in Metro Manila. If the post-ECQ infection rate is allowed to recover by half of the amount it decreased during the lockdown over a two-week period, then our model predicts that the total number of infected cases of COVID-19 will grow from 15,952 to 35,235 and that the total number of deaths will rise from 1,356 to 2,415 by August 1, 2020. However, it is important to emphasize that these increases are not inevitable. They can be offset with a rigorous tracking, testing, and tracing program that seeks to limit community spread by breaking chains of viral transmission (Hellewell et al., 2020).

**Figure 4:**
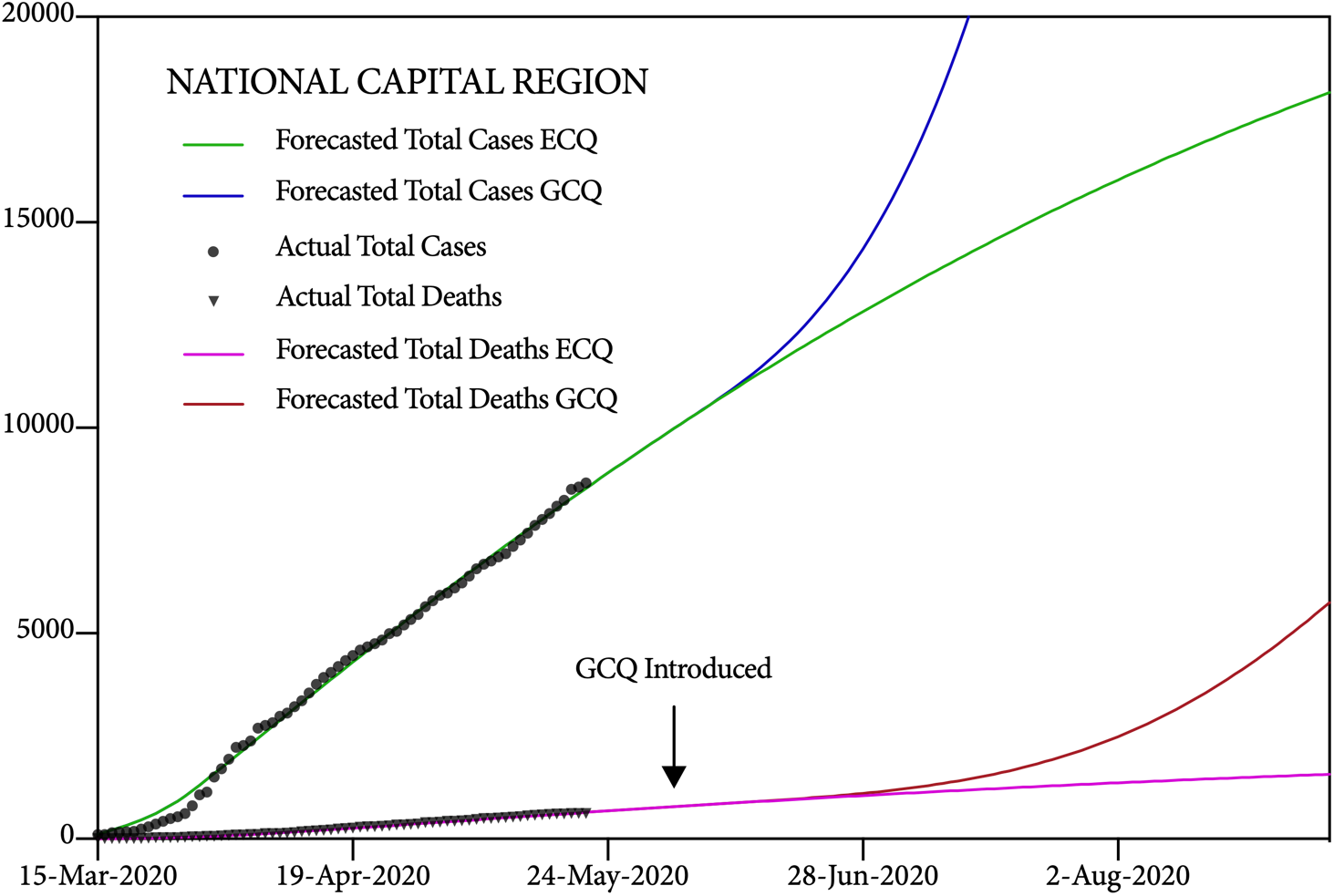
Modeling the Impact of the General Community Quarantine (GCQ) in Metro Manila from June 1, 2020. Forecasted estimates of total COVID-19 cases, total active cases, and total deaths for the National Capital Region for the ECQ and GCQ, compared to the data provided by the Department of Health of the Philippines.

Given the observation that typically, only 20% of a population contributes to at least 80% of the potential to transmit infectious disease (Endo et al., 2020; Woolhouse et al., 1997), we recommend that tracking, testing, and tracing programs in Metro Manila focus their efforts on limiting the impact of superspreading events (SSEs) which are associated with both explosive growth early in an outbreak and sustained transmission in later stages (Frieden & Lee, 2020).

How much testing and tracing capacity will be needed in Metro Manila to keep the pandemic at bay? A team of scholars at the Edmond J. Safra Center at Harvard University has recently published recommendations for communities of different sizes and at different stages in controlling their pandemics (Allen et al., 2020). They propose that every municipality with a moderate infection rate, i.e., with less than 1% prevalence of active virus in its population – for the most part, the NCR appears to fulfill this criterion – should hire sixty teams of five tracers for every death per day that they observe in their community, and maintain a testing capacity of 2,500 tests for every death per day.

On May 22, 2020, the seven-day running average of deaths per day recorded in Metro Manila was six deaths per day. This means that at this stage of the pandemic, according to the recommendations of the Harvard team, the NCR would need a testing capacity of 15,000 tests per day and 1,800 contact tracers working in call centers scattered throughout the region, to control its local pandemic. Geographic distribution of this testing and contact tracing capacity should correspond to the severity of the local pandemics in each of the cities of Metro Manila, with a particular focus on the City of Manila and Quezon City.

In sum, though they were necessary to squash the initial wave of the COVID-19 pandemic, community-wide lockdowns are not sustainable in the long run because of their grave socio-economic impact, especially on the lives of the poor (Nicola et al., 2020). Our modeling suggests that the government-imposed Enhanced Community Quarantine (ECQ) has successfully limited the spread of the pandemic in Metro Manila. It has given the Philippine government precious time to ramp up the social and medical infrastructure that will be needed to mitigate and contain the virus for the foreseeable future until a vaccine is invented. Resources must now be invested to increase testing and contact tracing capacity to prevent future outbreaks of COVID-19, and to increase the capacity of the Philippine health care system, including sufficient supplies of PPE, so that it will be able to deal with a possible second wave of the pandemic later this year.

## Data Availability

This epidemiological study used publicly available data sets provided by the Department of Health of the Philippines at https://www.doh.gov.ph/covid19tracker

## ACKNOWLEDGEMENTS

We are grateful to Daniel Luchangco, M.D., and Nicholas Matsakis, Ph.D., for helpful discussion, and to Dean Rey Donne S. Papa, Ph.D., of the College of Science of the University of Santo Tomas for his support of this project.

